# A multivariable Mendelian randomization to appraise the pleiotropy between intelligence, education, and bipolar disorder in relation to schizophrenia

**DOI:** 10.1101/19012401

**Authors:** Charleen D. Adams

## Abstract

Education and intelligence are highly correlated and inversely associated with schizophrenia. Counterintuitively, education genetically associates with an increased risk for the disease. To investigate why, this study applies a multivariable Mendelian randomization of intelligence and education. For those without college degrees, older age of finishing school associates with a decreased likelihood of schizophrenia—independent of intelligence—and, hence, may be entangled with the health inequalities reflecting differences in education. A different picture is observed for schooling years inclusive of college: more years of schooling increases the likelihood of schizophrenia, whereas higher intelligence distinctly and independently decreases it. This implies the pleiotropy between years of schooling and schizophrenia is horizontal and likely confounded by a third trait influencing education. A multivariable Mendelian randomization of schooling years and bipolar disorder reveals that the increased risk of schizophrenia conferred by more schooling years is an artefact of bipolar disorder – not education.

---

Schizophrenia is a heterogeneous neurological syndrome, typically presenting in early adolescence, and observationally associated with lower intelligence and lower educational attainment^1–3^.

Counterintuitively, education, which is positively associated with many health outcomes^4,5^, is genetically associated with an increased risk for schizophrenia^3^. Intelligence and education are highly positively correlated both phenotypically (r=0.8)^6^ and genetically (r=0.7)^7^. The traits are bidirectionally causally related: higher intelligence causes more years of schooling and more years of schooling increases intelligence^8^. The interwoven traits are also pleiotropically related to schizophrenia: a recent genome-wide association (GWA) study found evidence of an increased risk for schizophrenia for the single-nucleotide polymorphisms (SNPs) tagging years of schooling (*P* = 3.2 × 10^−4^) and strong genetic covariance between cognitive performance and increased years of schooling (*P* = 9.9 × 10^−50^)^9^.

Three possible explanations exist for the associations between intelligence, education, and schizophrenia: vertical, horizontal, and confounding pleiotropy (Fig. 1). Uncovering the nature of these relationships could inform interventional strategies. To that end, this study uses univariable and multivariable Mendelian randomization (MR) to appraise these pleiotropic relationships.

**Fig. 1.**
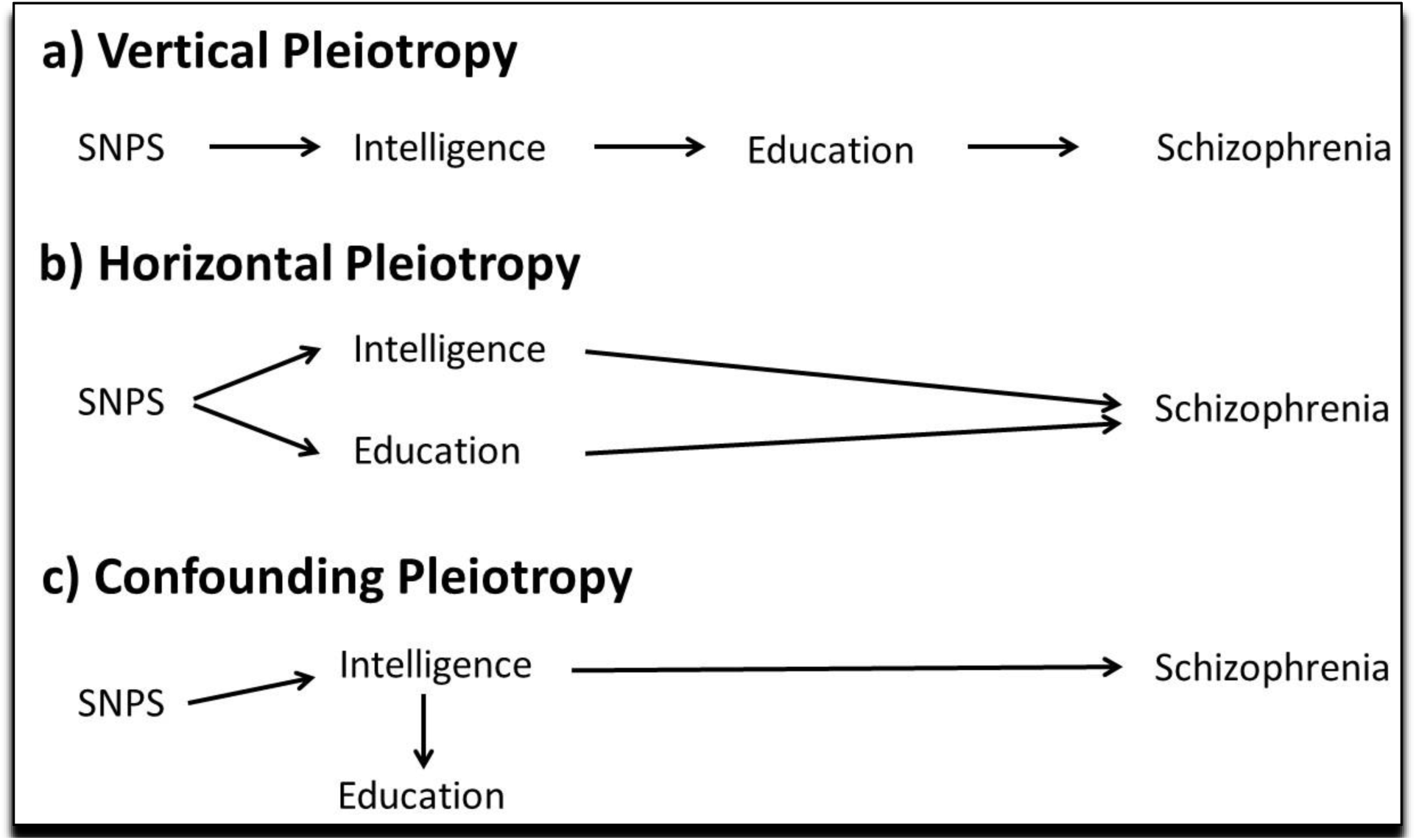
Possible explanations for the pleiotropy between intelligence, education and schizophrenia. An example of vertical pleiotropy would be the SNPs for intelligence influencing schizophrenia (only) through their effect on education. Vice versa, the SNPs for education might influence schizophrenia (only) through their effect on intelligence. Since education influences intelligence, an increase in intelligence from education might influence risk for schizophrenia (**a**). An example of horizontal pleiotropy would be if the SNPs for intelligence and/or the SNPs for education have independent, direct effects on schizophrenia (**b**). An example of confounding pleiotropy would be if education has no influence on schizophrenia but appears to due to strong association with IQ. Vice versa, IQ might not influence on schizophrenia but appears to due to strong association with education (**c**). Multivariate MR can be used to investigate these relationships.

The MR findings show that, for those without college degrees, older age of finishing school (Education Age) associates with a decreased likelihood of schizophrenia—independent of intelligence—and, hence, may be entangled with the health inequalities reflecting differences in education. As such, targeted strategies to retain at-risk adolescents in school may be both warranted as a prevention against schizophrenia and difficult to implement societally.

A different picture is observed for years of schooling inclusive of college (Education Years): more schooling years increases the likelihood of schizophrenia, whereas higher intelligence distinctly and independently decreases it. This implies the pleiotropy between schooling years and schizophrenia is horizontal and likely confounded by a third trait also influencing education. Further to this, bipolar disorder, associated observationally with both higher education and schizophrenia^3,10,11^, was investigated along with education, also using multivariable MR. The findings reveal that the increased risk of schizophrenia conferred by more schooling years is an artefact of bipolar disorder – not education.

## Results

### MR-Egger intercept

**Tables 1 and 2** contain the results for (i) the univariable (total) effects of education and intelligence on schizophrenia, (ii) the univariable results for the (total) effect of bipolar disorder on schizophrenia, and (iii) the bidirectional effects of education and intelligence. The MR-Egger intercept column is shaded grey, as its interpretation is different than that of the other tests; the MR-Egger intercept provides a test for directional pleiotropy and an assessment of the validity of the instrument assumptions^12^. If the intercept is not different than 1 on the exponentiated scale (or 0 on the non-exponentiated scale), that indicates a lack of evidence for bias in the IVW estimate, as is the case for all the univariable results.

**Table 1.**
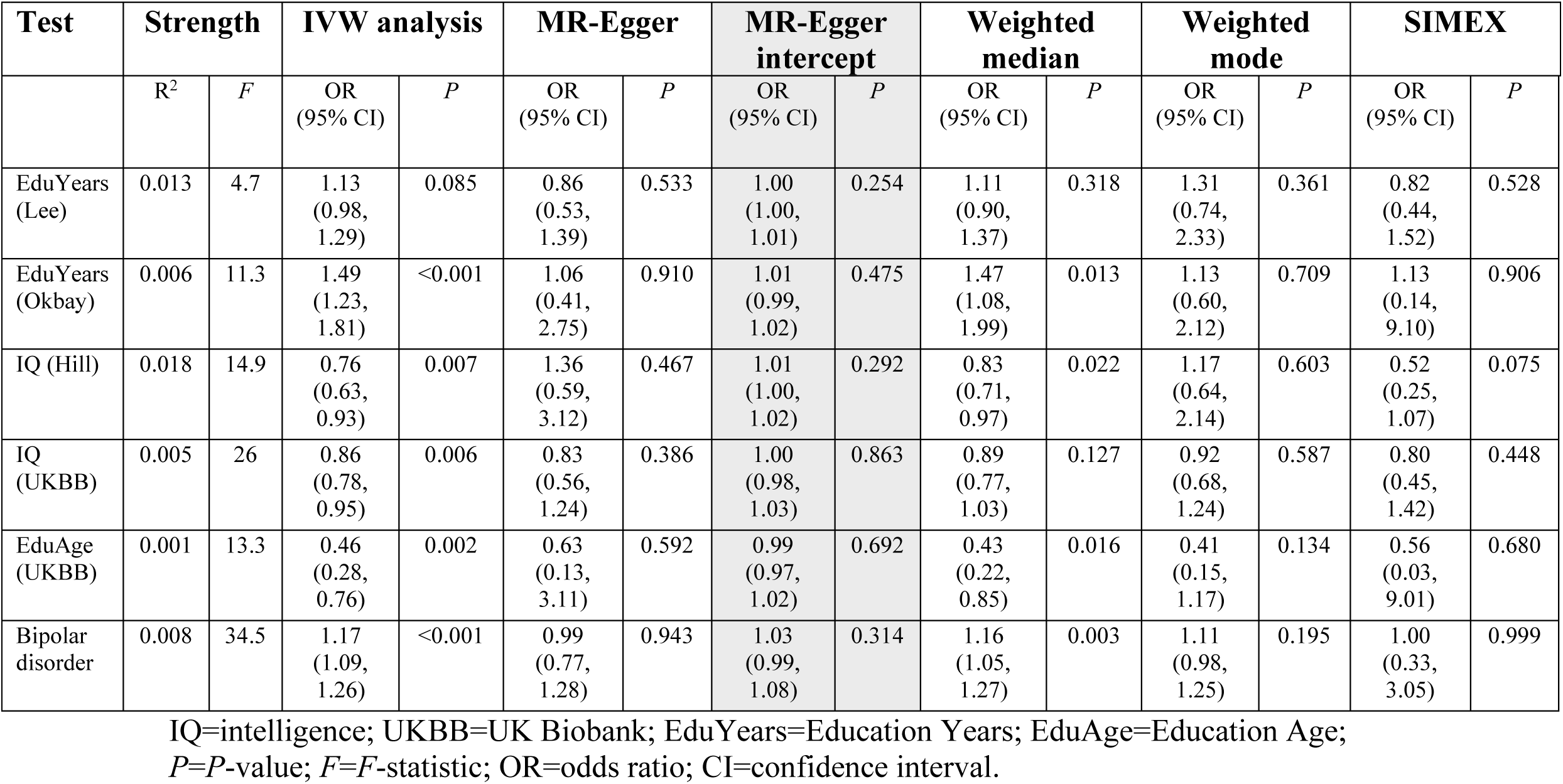
Univariable estimates of the effect of education, intelligence, and bipolar disorder on schizophrenia.

| Test | Strength |  | IVW analysis |  | MR-Egger |  | MR-Egger intercept |  | Weighted median |  | Weighted mode |  | SIMEX |  |
| --- | --- | --- | --- | --- | --- | --- | --- | --- | --- | --- | --- | --- | --- | --- |
|  | R<sup>2</sup> | F | OR (95% CI) | P | OR (95% CI) | P | OR (95% CI) | P | OR (95% CI) | P | OR (95% CI) | P | OR (95% CI) | P |
| EduYears (Lee) | 0.013 | 4.7 | 1.13 (0.98, 1.29) | 0.085 | 0.86 (0.53, 1.39) | 0.533 | 1.00 (1.00, 1.01) | 0.254 | 1.11 (0.90, 1.37) | 0.318 | 1.31 (0.74, 2.33) | 0.361 | 0.82 (0.44, 1.52) | 0.528 |
| EduYears (Okbay) | 0.006 | 11.3 | 1.49 (1.23, 1.81) | <0.001 | 1.06 (0.41, 2.75) | 0.910 | 1.01 (0.99, 1.02) | 0.475 | 1.47 (1.08, 1.99) | 0.013 | 1.13 (0.60, 2.12) | 0.709 | 1.13 (0.14, 9.10) | 0.906 |
| IQ (Hill) | 0.018 | 14.9 | 0.76 (0.63, 0.93) | 0.007 | 1.36 (0.59, 3.12) | 0.467 | 1.01 (1.00, 1.02) | 0.292 | 0.83 (0.71, 0.97) | 0.022 | 1.17 (0.64, 2.14) | 0.603 | 0.52 (0.25, 1.07) | 0.075 |
| IQ (UKBB) | 0.005 | 26 | 0.86 (0.78, 0.95) | 0.006 | 0.83 (0.56, 1.24) | 0.386 | 1.00 (0.98, 1.03) | 0.863 | 0.89 (0.77, 1.03) | 0.127 | 0.92 (0.68, 1.24) | 0.587 | 0.80 (0.45, 1.42) | 0.448 |
| EduAge (UKBB) | 0.001 | 13.3 | 0.46 (0.28, 0.76) | 0.002 | 0.63 (0.13, 3.11) | 0.592 | 0.99 (0.97, 1.02) | 0.692 | 0.43 (0.22, 0.85) | 0.016 | 0.41 (0.15, 1.17) | 0.134 | 0.56 (0.03, 9.01) | 0.680 |
| Bipolar disorder | 0.008 | 34.5 | 1.17 (1.09, 1.26) | <0.001 | 0.99 (0.77, 1.28) | 0.943 | 1.03 (0.99, 1.08) | 0.314 | 1.16 (1.05, 1.27) | 0.003 | 1.11 (0.98, 1.25) | 0.195 | 1.00 (0.33, 3.05) | 0.999 |

IQ=intelligence; UKBB=UK Biobank; EduYears=Education Years; EduAge=Education Age; $P$ = $P$ -value; $F$ = $F$ -statistic; OR=odds ratio; CI=confidence interval.

**Table 2.**
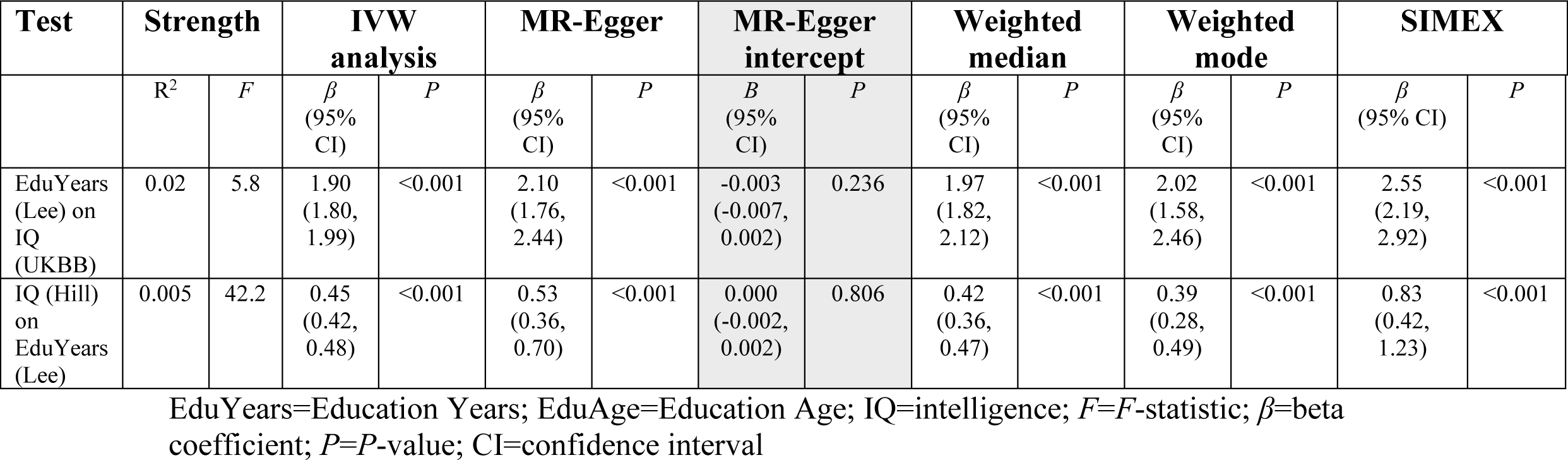
Bidirectional relationship between Education Years and intelligence.

### Education Years (Lee instrument) on schizophrenia

An increased (but null) effect on schizophrenia is observed for Education Years (odds ratio (OR) for schizophrenia per SD increase in years of schooling: IVW estimate 1.13; 95% CI 0.98, 1.29; *P* = 0.085). The sensitivity estimators are discrepant both in direction and magnitude of effects, indicating possible unwanted pleiotropy. SIMEX correction did not ameliorate this for the MR-Egger estimate.

### Education Years (Okbay instrument) on schizophrenia

In contrast, a robust increased risk for schizophrenia is observed for the Education Years: OR for schizophrenia per SD increase in Education Years: instrument estimate 1.49; 95% CI 1.23, 1.81; P<0.001). There is comportment in the direction of effects among the sensitivity estimators. The weak *F*-statistic for the Lee instrument may explain the discrepancy between the Lee and Okbay results.

### Education Age on schizophrenia

A strong protective effect against schizophrenia is observed for Education Age (OR for schizophrenia per SD increase in Education Age): IVW estimate 0.46; 95% CI 0.28, 0.76; *P*=0.002). The sensitivity estimators align both in direction and magnitude of effects.

### Intelligence (Hill instrument) on schizophrenia

A protective effect of intelligence against schizophrenia is observed for both the Hill and UK Biobank instrumental variables. There is, however, substantial disagreement between the IVW and MR-Egger estimates for the Hill instrument, which was rescued by SIMEX correction (the direction of the effect is reversed towards that of the IVW). The remaining discordance in the sensitivity estimators for the Hill instrument likely indicates pleiotropy: OR for schizophrenia per SD increase in intelligence: IVW estimate 0.76; 95% CI 0.63, 0.93; *P*=0.007.

### Intelligence (UK Biobank instrument) on schizophrenia

A robust protective effect against schizophrenia is observed for the UK Biobank instrument (OR for per SD increase in intelligence): IVW estimate 0.86; 95% CI 0.78, 0.95; *P*=0.006. The sensitivity estimators align.

### Bipolar disorder on schizophrenia

An increased risk for schizophrenia is observed per genetic liability to bipolar disorder (IVW estimate 1.17; 95% CI 1.09, 1.26; *P*<0.001). The effect estimate is reversed for the MR-Egger estimator, and the magnitudes of the various estimators vary, possibly indicative of some unwanted pleiotropy.

### Multivariable results

Fig. 2 contains the comparison of the univariable and multivariable (adjusted) estimates for the effects of education and intelligence, and bipolar disorder and education on schizophrenia.

**Fig. 2.**
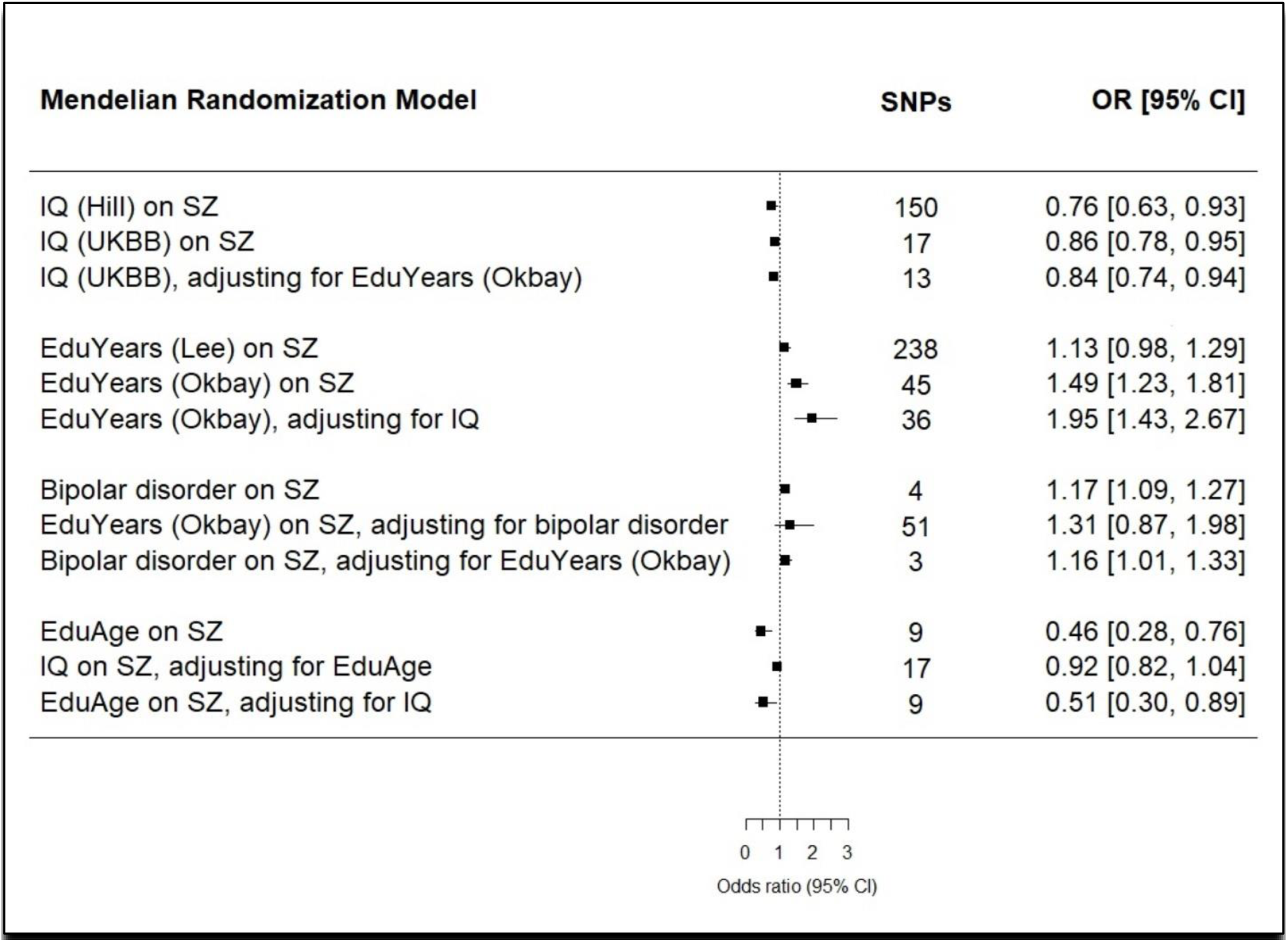
Comparison of univariable and multivariable (adjusted) estimates of the effects of education and intelligence and bipolar disorder and education on schizophrenia. IQ=intelligence; EduYears=Education Years; EduAge=Education Age; SZ=schizophrenia; UKBB=UK Biobank; OR=odds ratio; CI=confidence interval.

### Intelligence, adjusting for Education Age

The impact of intelligence on schizophrenia attenuates to the null when adjusting for Education Age (adjusted OR for schizophrenia per SD increase in intelligence: IVW estimate 0.92; 0.82, 1.04; *P*=0.219). One explanation for the difference observed between the univariable and multivariable MR estimates for the effect of intelligence on schizophrenia is that intelligence affects schizophrenia through its effect on Education Age, rather than through a direct effect on schizophrenia.

### Intelligence, adjusting for Education Years

The protective effect of intelligence remains after adjusting for Education Years (adjusted OR for schizophrenia per SD increase in intelligence: IVW estimate 0.84; 95% CI 0.74, 0.94). This suggests intelligence has a robust and direct protective effect against schizophrenia. The effect attenuates some in comparison to the univariable model, perhaps reflecting the loss of the contribution of Education Years to intelligence (Table 3 and Fig. 7).

**Table 3.** Univariable, multivariable, and bidirectional Mendelian randomization models.

| Type | Mendelian randomization model (source GWAS) | SNPs |
| --- | --- | --- |
| Univariable | Intelligence (Hill) on schizophrenia | 150 |
| Univariable | Intelligence (UK Biobank) on schizophrenia | 17 |
| Multivariable | Intelligence (UK Biobank), adjusting for Education Years (Okbay) | 13 |
| Univariable | Education Years (Lee) on schizophrenia | 238 |
| Univariable | Education Years (Okbay) on schizophrenia | 45 |
| Multivariable | Education Years (Okbay), adjusting for intelligence (UK Biobank) | 36 |
| Univariable | Education Age (UK Biobank) on schizophrenia | 9 |
| Multivariable | Intelligence (UK Biobank) on schizophrenia, adjusting for Education Age (UK Biobank) | 17 |
| Multivariable | Education Age (UK Biobank) on schizophrenia, adjusting for intelligence (UK Biobank) | 9 |
| Bidirection 1 | Intelligence (Hill) on Education Years (Lee) | 35 |
| Bidirection 2 | Education Years (Lee) on intelligence (UK Biobank) | 299 |
| Multivariable | Education Years (Okbay) on schizophrenia, adjusting for bipolar disorder | 51 |
| Multivariable | Bipolar disorder on schizophrenia, adjusting for Education Years | 3 |
| Univariable | Bipolar disorder on schizophrenia | 4 |

### Education Age, adjusting for intelligence

A direct protective effect against schizophrenia is observed for Education Age (adjusted OR for schizophrenia per SD increase in Education Age: IVW estimate 0.51; 95% 0.30, 0.89; *P*=0.02).

### Education Years, adjusting for intelligence

An increased risk for schizophrenia is observed for Education Years (adjusted OR for schizophrenia per SD increase in Education Years: IVW estimate 1.95; 95% 1.43, 2.67; *P*<0.001). Together with the multivariable results for intelligence when adjusted for Education Years, these findings strongly suggest that the underlying pleiotropy between intelligence and Education Years is horizontal in relationship to schizophrenia (Fig. 1) and that the relationship is additionally caught up by the presence of an unmeasured confounder. The horizontal pleiotropy and opposing directions of effect for education and intelligence prompted a univariable investigation of bipolar disorder and schizophrenia and a multivariable Mendelian randomization of bipolar disorder and education on schizophrenia. The proposed hypothesis is seen in Fig. 3.

**Fig. 3.**
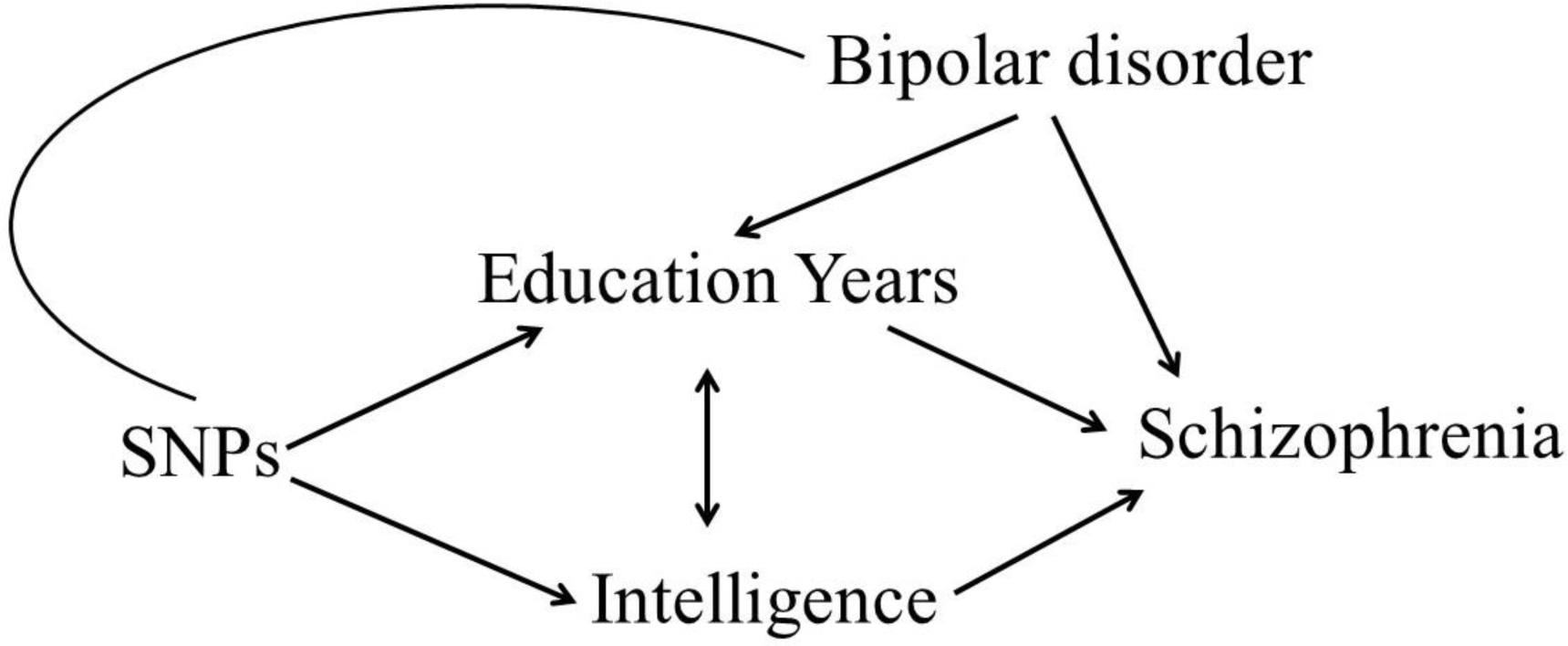
Hypothesized relationships between Education Years, intelligence, bipolar disorder, and schizophrenia suggested by the multivariable analysis of education and intelligence on schizophrenia. DAG=directed acyclic graph.

### Education Years on schizophrenia, adjusting for bipolar disorder

The increased risk for Education Years on schizophrenia attenuated to the null when accounting for bipolar disorder (adjusted OR: IVW estimate 1.31; 95% CI 0.87, 1.98; *P*=0.207).

### Bipolar disorder on schizophrenia, adjusting for Education Years

A direct, increased risk is observed for genetic liability to bipolar disorder on schizophrenia (adjusted OR for schizophrenia: IVW estimate 1.16, 95% CI 1.01, 1.33; *P*=0.033).

### Bidirectional relationship between Education Years and intelligence

Table 2 and Fig. 4 depict the results for the bidirectional analysis of Education Years and intelligence. A SD-unit higher intelligence causes more Education Years (*β* 0.45, 95% CI 0.42, 0.48; *P*<0.001) and a SD-year more of Education Years increases intelligence (*β* 1.90, 95% CI 1.80, 1.99; *P*<0.001). These findings replicate those of Anderson *et al*. (2018)^8^.

**Fig. 4.**
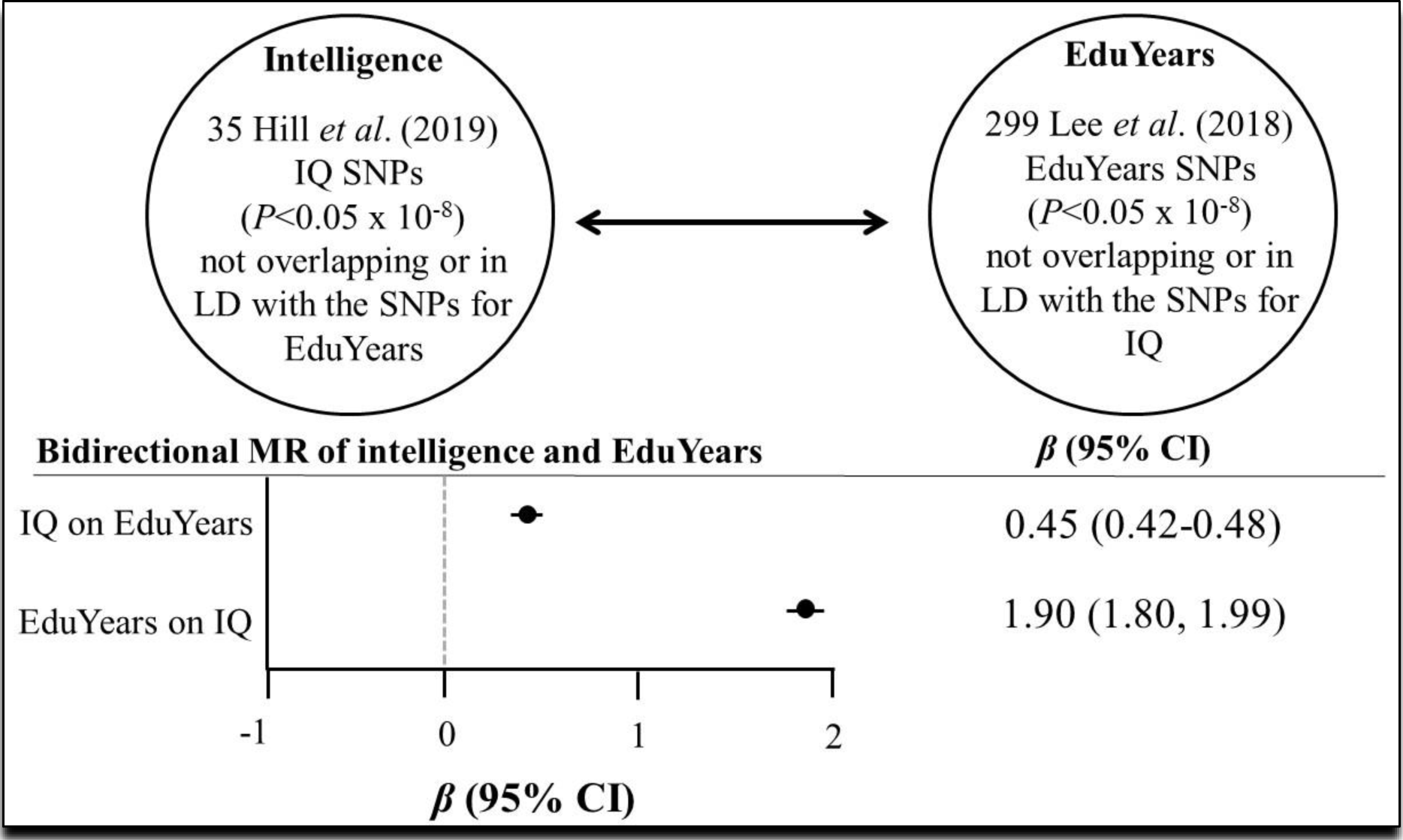
Bidirectional relationship between intelligence (IQ) and Education Years (EduYears). LD=linkage disequilibrium.

## Discussion

Educational attainment has been described as feature of the bipolar disorder^10,11^. Bipolar disorder shares some cognitive deficits and genetic overlap with schizophrenia, but also predisposes to cognitive adeptness and creativity that distinguish it from the more neurodevelopmental aspects of schizophrenia^3^. This complex picture is reflected in the horizontal and confounding pleiotropy uncovered by the multivariate analyses here. Specifically, when bipolar disorder is not accounted for, it appears that more years of schooling increase risk for schizophrenia. Hence, bipolar disorder is a confounder of the relationship between education and schizophrenia. Since more years of schooling increase intelligence and higher intelligence strongly protects against schizophrenia, these findings imply that staying in school is neuroprotective.

For those without college degrees, education—not intelligence—acts as the mechanism conferring protection. The implications of this are uncertain, since the protective effect is likely to be entangled with the social inequalities linked to educational attainment. Nonetheless, efforts to retain at-risk adolescents in school, especially those beginning to show features of cognitive impairment, may be worth exploring.

The primary strength of this study is that it capitalizes on the power of seven large GWA studies to probe these complexly related traits. It is the most detailed and comprehensive joint investigation of them to date. An unintended benefit of doing so demonstrates the value of these massive public datasets for etiologic discovery.

The study has several limitations. MR critically relies on the validity of the instrumental variables. As such, measures were taken to assess the robustness of the analyses to potential unwanted pleiotropy, including the use of instruments lacking between-SNP heterogeneity and comparison of the IVW estimate with a battery of sensitivity estimators, each making different assumptions.

Another possible limitation, which, like unwanted pleiotropy, cannot be entirely ruled out, is the possible introduction of bias caused by some instances of the same individuals being included in the GWA studies of both the exposures and the outcomes. The greatest overlap is likely to be for the Lee Education Years instrument on intelligence and the Hill intelligence instrument on Lee’s Education Years. However, since that bidirectional appraisal is a replication of Anderson *et al*.’s (2018) study that used non-overlapping samples with comparable results, the impact of the bias is likely to be minimal.

## Methods

### Conceptual approach

MR is an instrumental variables technique, analogous to a randomized control trial. It capitalizes on several features of the genome for causal inference: 1) the random assortment of alleles (Mendel’s Laws of Inheritance dictating that alleles segregate randomly from parents to offspring) and 2) pleiotropy (genes influencing more than one trait)^13–15^. Two-sample MR (Fig. 5) uses summary statistics from two genome-wide association (GWA) studies^12,16–20^, and multivariable MR is a further extension of the procedure that permits adjustment^21^.

**Fig. 5.**
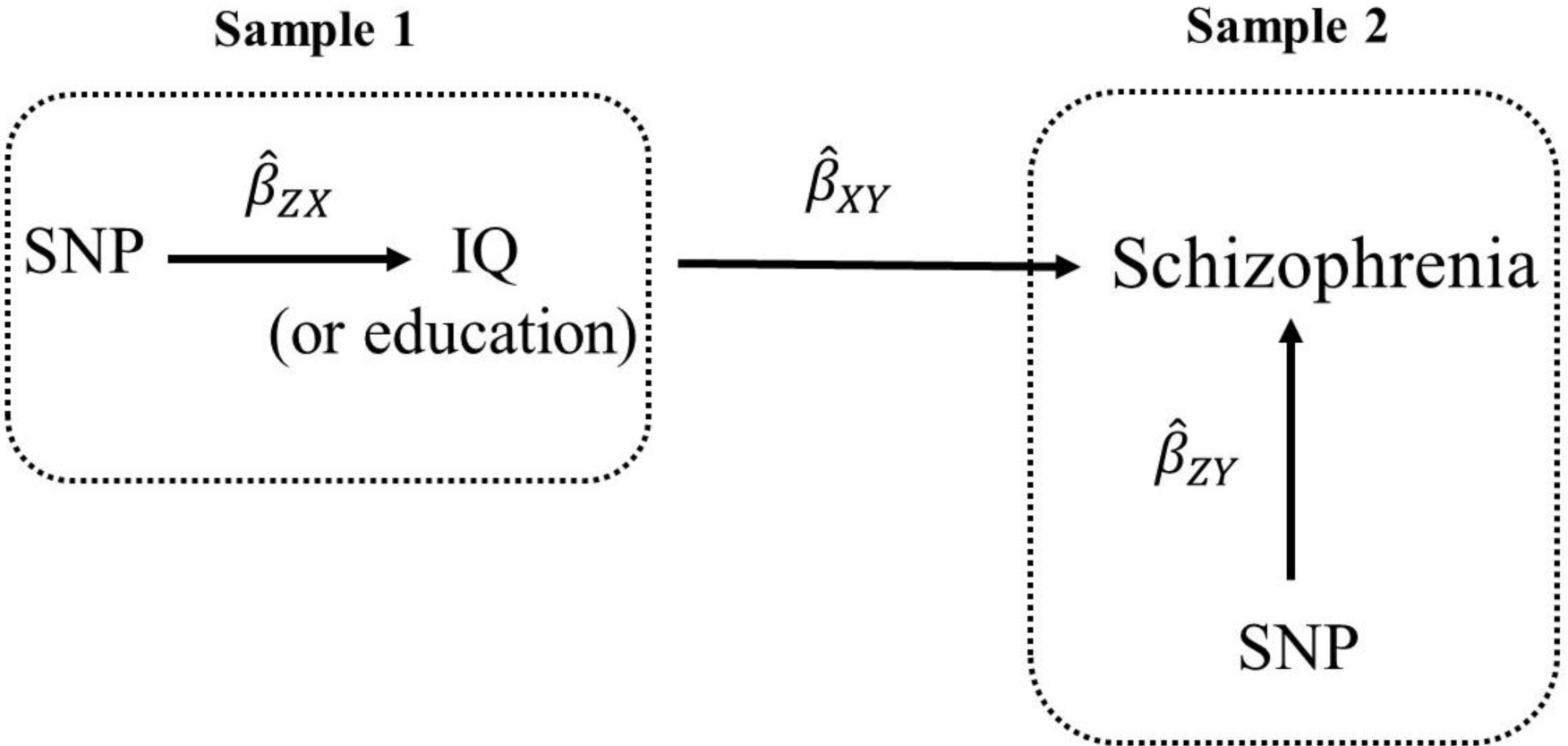
Two-sample Mendelian randomization testing the causal effect of intelligence or education on schizophrenia. Estimates of the SNP-intelligence (or SNP-education) associations 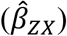 are calculated in sample 1 (from GWA study of intelligence or GWA study of education). The association between these same SNPs and schizophrenia are then estimated in sample 2 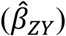 (from a schizophrenia GWA study). These estimates are combined into Wald ratios 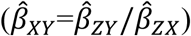. The 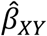 estimates are meta-analyzed using the inverse-variance weighted analysis 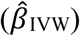 method. The IVW method produces an overall causal estimate of intelligence and/or education on schizophrenia.

### Mendelian randomization assumptions

In order for MR to be valid, three assumptions must hold: (i) the SNPs acting as the instrumental variables must be strongly associated with the exposure; (ii) the instrumental variables must be independent of confounders of the exposure and the outcome; and (iii) the instrumental variables must be associated with the outcome only through the exposure^18,22^. When violated, assumption (iii) describes horizontal (Fig. 1b) pleiotropy, which can invalidate causal inference from vertical (Fig. 1a) pleiotropy probed in univariable MR designs.

### GWA study data sources for instruments

#### Education Age on schizophrenia

Two measures of education were selected to instrument education: age at completion of full-time schooling without a college degree (Education Age) and years of schooling inclusive of college (Education Years). The Education Age measure was obtained from field 845 in the UK Biobank project^23,24^. Participants were asked if they had a college or university degree. Those without a college or university degree were asked what age they left continuous full-time education. Summary statistics for a GWA study of Education Age (adjusted for sex and 10 principal components), including 226,899 UK Biobank participants who answered field 845, are publicly available; the GWA study was performed by the Neale lab, after transforming the item into a normally distributed quantitative variable^25^ (SNP coefficients per standard deviation (SD) units of Education Age). Because the instrument for Education Age captures only those without college or university degrees, the inference from the use of Education Age as an instrument is restricted to without college or university degrees.

The *F*-statistic, a function of how much variance in a trait is explained by an instrument (R^2^), the sample size, and the number of SNPs in an instrument, provides an indication of instrument strength^26^. *F*-statistics <10 are conventionally considered to be weak^27^. The *F*-statistic for the Education Age instrument is 13.3.

#### Education Years on schizophrenia

The primary years of schooling measure was obtained from the Lee *et al*. (2018) GWA study of 1,131,881 participants of European ancestry from 71 cohorts ^28^. Education Years was measured for those who were at least 30 years of age, and International Standard Classification of Education (ISCED) categories were used to impute a years-of-education equivalent (SNP coefficients per SD units of years of schooling). The *F*-statistic for the Lee Education Years instrument is 4.7, indicating the instrument may be weak. Due to this, a second measure of Education Years from a smaller GWA study of years of schooling was used to construct a second instrument for Education Years^9^. The Okbay et al. (2016) GWA study used the same construction of Education Years as did Lee *et al*. (2018) GWA study and contained 293,723 participants of European ancestry^9^. The Okbay Education Years instrument has an *F*-statistic of 11.3. Because it is aptly strong, the Okbay Education Years instrument was used in the multivariate model of intelligence and education on schizophrenia.

#### Education Years on intelligence (Bidirection 1)

The Lee *et al*. (2018) GWA study was used to extract SNPs for the first part of the bidirectional analysis of education on intelligence. The instrument has an *F*-statistic of 5.8, indicating it may be inadequately strong. However, a bidirectional appraisal of Education Years and intelligence using the Okbay *et al*. (2016) GWA study for instrumental variables was previously reported^8^. The Anderson *et al*. study is treated as a natural-history sensitivity analysis, since they included fewer and (likely) stronger SNPs (148 compared to 299, respectively), which can increase the *F* parameter^29^. (See Table 3 for a list of the number of selected SNPs for each of the instrumental variables).

#### Intelligence on schizophrenia (Hill instrument)

Two GWA studies were used to create instruments for intelligence. The first came from the Hill *et al*. (2019), which included 248,482 individuals of European ancestry (SNP coefficients per one SD increase in intelligence test scores^7^. The instrument’s *F*-statistic is 14.9.

#### Intelligence on schizophrenia (UK Biobank instrument)

A second instrument for intelligence was constructed from a GWA study performed by the Neale lab using the UK Biobank measure for fluid intelligence (field 20016) (n=108,818). The participants answered 13 logic questions within two minutes and the number of correct answers were summed. The data were transformed into a normally distributed quantitative variable (SNP coefficients per one SD unit increase in fluid intelligence score)^25^. The instrument’s *F*-statistic is 26.

#### Intelligence on education (Bidirection 2)

The Hill *et al*. (2019) GWA study of intelligence was used for the second part of the bidirectional analysis of intelligence and education. The instrument has an *F*-statistic of 42.2.

#### Bipolar disorder on schizophrenia

A GWA of bipolar disorder containing 16,731 participants of European descent (of which 7,481 were cases) was available for the instrument for bipolar disorder^30^. The instrument has an *F*-statistic of 34.5.

### GWA study data sources for outcomes

#### Intelligence

Because the full GWA study summary data were unavailable for the Hill GWA study of intelligence, the UK Biobank GWA study of intelligence (n=108,818) was used as the outcome GWA study for the tests of Education Years and Education Age on intelligence.

#### Education (Education Years)

Full summary data were available for 766,345 participants in the Lee *et al*. Education Years GWAS.

#### Education (Education Age)

Full summary data were available for 226,899 participants in the UK Biobank Education Age GWAS.

#### Schizophrenia

Full summary data were available for a schizophrenia GWA study dataset containing 82,315 participants of European ancestry, of which 35,476 were cases^31^.

#### Instrument construction

For each instrument 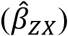, independent (those not in linkage disequilibrium, LD; R^2^ < 0.01) single-nucleotide polymorphisms (SNPs) associated at genome-wide significance (P < 5 x 10^-8^) with a trait were extracted from within their respective GWA study. The summary statistics for the instrument-associated SNPs were then extracted from an outcome GWA study 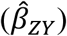. SNP-exposure and SNP-outcome associations were harmonized with the “harmonization_data” function within the MR-Base “TwoSampleMendelian randomization” package within R^16^. Harmonized SNP-exposure and SNP-outcome associations were combined with the IVW method (Fig. 2).

For the bidirectional associations between intelligence and schooling years, SNPs tagging both traits at genome-wide significance and/or SNPs that were in LD between intelligence and schooling years were excluded. This is because overlapping SNPs can invalidate bidirectional MR findings^20^. In addition, for all instrumental variables, RadialMR regression^32^ was run to detect SNP outliers. Outlier SNPs were removed. All instrumental variables included in this analysis have Cochrane’s *Q*-statistic *P*-values indicating no evidence for heterogeneity between SNPs^33^ (heterogeneity statistics are provided in Supplementary Tables 4, 7, 10, 13, 16, 21, 24, 27, and 30).

#### Sensitivity analyses

To address possible violations to MR assumption (iii), MR-Egger regression, weighted median, and weighted mode MR methods were run as complements to the IVW method for the univariable models. When the magnitudes and directions of the various MR methods comport across estimators, this lack of heterogeneity is a screen against pleiotropy. The reason for this is that various MR sensitivity estimators make different assumptions about the underlying nature of pleiotropy. It is unlikely there would be homogeneity in the direction and magnitudes of their effect estimates if there were substantial violations to the pleiotropy assumption. An extensive description of the different MR methods and the different assumptions they make about pleiotropy have been previously reported^34–36^. In addition, a SIMEX correction was performed for all univariate tests to correct potentional regression to the null in the MR-Egger estimates^37^ (Supplementary Tables 5, 8, 11, 14, 17, 22, 25, 28, 31).

#### Number of tests

In total, 14 MR tests were run. Table 3 contains a list of the tests and the number of instrumental variables (detailed characteristics for the individual SNPs used in each model are provided in Supplementary Tables 3, 6, 9, 12, 15, 20, 23, 26, and 29). These 14 tests are not independent; a false-discovery rate (FDR)-correction was applied to the raw *P-*values to assess whether the penalization changed the inference (Supplementary Table 2). As it did not, the raw *P*-values are reported for the following reasons: the inference remained unchanged, the FDR-adjustment is overly conservative in this case, and *P*-values alone are not the best guide for causal inference^38^.

#### Statistical software

SIMEX corrections were perfomed in Stata SE/16.0. All other described analyses were performed in R version 3.5.2.

## Data Availability

All data is publicly available.

http://www.mrbase.org/

## Data availability

All data sources used for SNP-exposure and SNP-outcome associations are publicly available. The data for the Hill intelligence^7^ and Lee Education Years^28^ instruments were obtained directly from the supplementary files accompanying their primary papers. The remaining data used for these analyses are accessible within MR-Base http://www.mrbase.org/^16^.

## Code availability

A copy of the code used in this analysis is available at

## Acknowledgements

Thank you to the cohorts that made their GWA study summary data public.

## Additional information

Supplementary information accompanies this paper

